# Assessing synthetic data generation utility for cohort data secondary use

**DOI:** 10.1101/2025.06.09.25329247

**Authors:** Mattia Mazzoli, Janis Elfert, Paolo Sacerdoti, Marco Hirsch, Michael Davis Tira, Daniela Paolotti

## Abstract

Access to cohort data is critical for reproducing studies, validating hypotheses in new settings and testing new approaches with past data in light of new findings. Nevertheless, previously collected data are often inaccessible due to confidentiality and privacy standards. In recent years, thanks to the development of AI, the field of synthetic data generation has emerged as a possible solution to facilitate cohort data sharing. However, synthetic data generation faces a tradeoff between statistical fidelity and information disclosure, which becomes even more restrictive for high dimensionality data.

Here we assess the feasibility of employing state-of-the-art data synthesization and anonymization techniques to generate high-fidelity and privacy preserving synthetic datasets capable of meaningfully reproducing study results, thus benchmarking the potential of synthetic cohort data for public sharing.

We design a protocol relying on four public packages, seven privacy metrics and five synthesization algorithms. We employ data collected within the framework of the Verdi project (Influweb) and publicly available hospitalisation cohort datasets (MIMIC-III) to assess privacy levels preserved by multiple data synthetization algorithms in three different studies. We employ multiple privacy metrics to ensure that statistical fidelity in our datasets do not come at the cost of information disclosure. Finally, we qualitatively compare the similarity of results reproduced with the synthetic datasets, noting that determinants of health-related behavior and in-hospital mortality remain largely unchanged with respect to estimates performed on the original data.

Our findings show that synthetic data generation is a promising technique for public data sharing and study reproducibility. However, its broad application for new exploratory studies on complex or rare patterns may introduce limitations and biases, as fidelity of previously unobserved statistical relationships is not guaranteed.

## Introduction

### Data readiness

Health data readiness is fundamental to pandemic preparedness as it shapes our capacity to brace and respond to health threats and inform effective decision-making^1,2^. Data accessibility and sustainability, intended as the capacity to endure in time, are crucial to build preparedness as they enable the testing of novel hypotheses, treatments, intervention strategies, support the reproduction of past studies and grant continuous data streams for future analyses.

Collecting new data, e.g. in prospective cohort studies or clinical trials for drug testing, is costly both in terms of funding and personnel resources. The availability of previously collected data can boost research both for new hypotheses testing or study reproducibility, including simulations of clinical trials^3^. New studies may need to access and utilize health records previously collected by other entities to test reproducibility, for training, for comparison or for gaining deeper insights. Unfortunately, access to previously collected data is usually restricted by consent requirements if the new research falls outside of participants’ consent statement, or by privacy if the analysis requires access to individual level information, or by confidentiality if no other institution rather than the collector can access the data. While peculiar contexts may allow Independent Review Boards (IRB) and Ethics Committees to circumvent these constraints and allow studies to follow an expedited review, e.g. involving minimal risk data, a possible way of overcoming these limitations is represented by Synthetic Data Generation (SDG).

### SDG and open challenges

SDG aims at emulating sensitive input data whose disclosure is forbidden due to privacy risks or confidentiality requirements. A good SDG algorithm should preserve three aspects: utility, fidelity and privacy. Utility is case dependent and is defined by the scope of the data usage. It encodes the study purpose, i.e. the scope of the generated data, like estimating determinants of behavior, efficacy of treatment or analysing temporal trends. Fidelity represents the statistical similarity between the original and the synthetic dataset in a very general and multifaceted way. Fidelity can be assessed in terms of pairwise correlations but also higher-order and non-linear interactions between variables, similarity measures, clustering or other aspects related to spatial and temporal properties of the data, which may be disease-specific but critical in clinical data modeling. Fidelity is intrinsically determined by utility: the scope of the study defines the critical dimensions whose statistical similarity has to be preserved; the remaining features in the dataset can be neglected. Privacy is a wide aspect that represents the level of information that the algorithm or the generated data disclose from the original data, as presence, identity and sensitive attributes of individuals. The theoretical advantage of SDG is the sampling of individual-level artificial datasets from data generation models trained on the original dataset. The original data is never shared with the final user, hence removing any link between the synthetic data and real identities. SDG aims at preserving utility while maintaining high privacy levels and overcoming the vulnerability of classic anonymization models, which are exposed to linkage attacks, i.e. the re-identification of identities within the anonymous dataset. However, in practice SDG should be handled with care when applied to high-dimensional data since it does not offer complete safety from linkage attacks if outliers are reproduced or original records are leaked into the synthetic data, even if differential privacy algorithms were employed^4,5^.

SDG is a very suitable technique for healthcare research as it allows analysts and modelers to apply the same methodological pipelines employed on the original data, as data structure is preserved. Applications in healthcare are several^6,7^ and range from simulation and prediction of disease outcomes^8,9^, to testing and improvement of algorithms performance for outbreak detection^10^, estimate public health measures effectiveness^11^, development of EHR management software, education and training^12^. SDG opens the doors to public sharing and re-usability of data coming from individual surveys, hospital admissions and clinical trials^13^, by building data-readiness. Moreover, it broadens the breadth of research by providing the community with a wider diversity of datasets collected in different periods and settings. Following this opportunity, some institutions have already shared synthetic datasets of patients’ records, as the England’s National Health Service released three different synthetic datasets regarding Hospital Episodes^14^, the International Organization for Migration released synthetic datasets on human trafficking^15^, the Centers for Medicare and Medicaid Services released a synthetic dataset on millions of health insurance claims and drugs prescriptions^16^, RTI released the US synthetic household population^17^.

Despite being a field active for 30 years^18^, SDG still lacks recognized benchmark metrics for evaluating fidelity and privacy of synthetic datasets and relies on common metrics developed for anonymization techniques. While recent studies proposed SDG specific metrics for privacy evaluation like Targeted Correct Attribution Probability (TCAP)^19,20^, it is hard to read a consensus in the community over a set of golden standard metrics. Most anonymization metrics have been developed to preserve privacy while maintaining utility for low-dimensional datasets, whereas high dimensional data anonymization is very hard or even impracticable^4,21^.

The field is mainly driven by the advancement of the AI sector, which makes it a field in rapid evolution. Even today, there is still an ongoing debate within the community to define what are the best algorithms to tradeoff between fidelity and privacy, but most importantly, it is still unclear whether SDG may perform better than standard anonymization techniques, since its high fidelity comes at the expense of information disclosure^5^.

### The legal framework

Anonymization and pseudonymisation are defined and regulated by legislation on human research in health, both in the EU GDPR^22^ in Recital 26^23^ and the US HIPAA^24^ and 45CFR46^25^, however, even if SDG is kin to anonymization, it is never addressed explicitly by law. In the Article 29 Data Protection Working Party Opinion^26^ the EU Commission advisory body sets three tests that all anonymization techniques should satisfy to ensure appropriate privacy levels. Anonymization should ensure that no attacker would be able to isolate some or all attributes associated with an individual (outliers detection), nor link the information they already have in order to infer the presence of the individual in the records (membership disclosure), nor infer individuals’ information from the rest of their features (attribute disclosure).

### Privacy breach scenarios

When dealing with sensible individual data, like electronic health records (EHR), state-of-the-art privacy models aim at preventing worst-case scenarios involving attackers who have access to, or part of, the original data and know what algorithm was used to synthesize the data (white box attack). Adversaries may link the presence of an identity to an individual in the original data (membership inference), retrieve further information on users from whom they already have partial information (attribute disclosure), re-identify data (re-identification) solely on the basis of indirect identifiers (quasi-identifiers) or re-identify users using multiple releases of the original dataset (composition attack)^27^. Membership inference or attribute disclosure are linkage attacks and in these cases are powerful threats to privacy that need to be addressed.

### A second life for Verdi cohorts

Here we assess the feasibility and the advantage of employing SDG algorithms to allow reproducibility of past studies for cohorts data collected over the last three years by partners of the Verdi project consortium.

Here we rely on two datasets coming from previous data collection, one is the Influweb cohort data on influenza-like-illness (ILI) regarding patients’ symptoms and behavior and the second is the MIMIC-III database on vital signs, demographics, mortality and diagnoses of hospitalized patients. We employ state-of-the-art SDG algorithms to generate synthetic versions of the original datasets, test reproducibility of previous analyses from the literature and assess privacy of resulting datasets. Finally, we discuss whether we reach a satisfactory tradeoff between utility and privacy that allows us to share previously collected data in a way that maximizes reproducibility, favors new analyses and preserves privacy.

Our work has important implications since the Verdi project has been an important international initiative at the European level that contributed substantially to the understanding of the pandemic impact on individuals’ health and behavior. Therefore, the sharing of the data collected during this period would pave the way for further similar initiatives to follow and enhance data readiness.

## Methods

### Basic definitions

- **Quasi-identifier**: a quasi-identifier (QI) is a feature that typically relates to a group of people known to a particular group or the general public, e.g. age, gender, first name, date of birth. QIs combinations can help reveal the identity of individuals from a dataset.
- **Identifier**: it consists of a unique identifier that points to the identity of a person in the dataset. It is typically replaced by a hashed-id string in the so-called pseudonymization.
- **Sensitive attribute**: a sensitive attribute (SA) refers to information that must not be disclosed such as personal income or diagnoses.
- **Equivalence class**: an equivalence class (EC) is the subset of individuals with the same set of features, typically quasi-identifiers.

### Privacy metrics

Since the aim is to prevent as many attacks as possible and there are many different attack scenarios, several privacy metrics should be deployed, with each aiming to minimize the risk of vulnerabilities as much as possible. Below is a list of metrics that we employ in this work:

- **Differential Privacy**: Differential Privacy (DP) is a context-independent feature of the algorithm, rather than a feature of the data itself. A DP-process ensures that when synthesizing a given original dataset of individuals and a copy dataset obtained by excluding a given individual, the probability of observing a given feature in the two output datasets will not differ by more than *e^ε^*. High values of privacy loss, *ε*, may favor the attacker’s capacity of identifying the missing individual from the original data^28^. <u>Scenario</u>: assume an attacker who has complete knowledge on a dataset of students of a given school, e.g. the teacher. Among the students, one has an average mark of 5.5. The school wants to publish the list of students who have an average mark of 6 or above, but without letting anybody know who is missing. If proper differential privacy is not implemented^29^, the attacker will assess who the missing student is and retrieve sensitive information from the public dataset. <u>Purpose:</u> it contrasts membership inference and composition attacks^21^. The metric is imposed using *synthcity*.
- **TCAP**: Targeted Correct Attribution Probability (TCAP) assesses a white box attacker’s probability of correctly identifying an individual (or a SA) by knowing that the individual is in the dataset and by knowing part of the individuals’ features in the input data^19,20^. TCAP ranges from 0 (no risk) to 1 (maximum risk of disclosure). <u>Scenario</u>: assume that an attacker knows what algorithm has been used to generate the synthetic dataset, e.g. an ex-colleague. The attacker wants to re-identify an individual present in the original dataset using the synthetic data features. The attacker trains a machine learning algorithm to predict the users IDs of the original data from their features and tests it on the synthetic data. If TCAP is low enough, the attacker will have a very low probability of re-identifying users from synthetic data. <u>Purpose:</u> it contrasts attribute disclosure in white box attacks. The metric is computed using the package *SynthGauge*.
- **k-anonymity**: it describes the minimum size of ECs based on QIs in the dataset. If given the rarest individual in the dataset there are another 9 individuals with the same set of QIs, the dataset is said to be k=10 anonymous, meaning that an attacker will have a chance of 1/10 to guess an individual who matches that description^30^. The higher the *k*, the higher the privacy preserved, benchmark thresholds are set for *k = 5*. <u>Scenario</u>: assume an attacker who knows all the features of an individual and wants to know if this appears in a dataset of diabetes patients. If the dataset is 2-anonymous, i.e. there are two patients sharing the same set of QIs, the attacker will have a 50% chance of identifying the victim in the dataset, and potentially access more attributes. <u>Purpose</u>: it contrasts identity disclosure by maximising the size of the rarest EC based on QIs. The metric is computed using *pyCANON* and *Anjana*.
- **l-diversity**: L-diversity ensures a minimum level of diversity in the dataset by granting that each SA has at least *l* different values in each EC^31^, e.g. it avoids the scenario in which all individuals of one age class have the same diagnosed disease. <u>Scenario</u>: assume that the attacker has access to a dataset of cancer patients for which the SA is “has cancer”. The attacker knows that the individual is in the dataset and knows the victim’s location of residence is Rome. If all patients reporting Rome as location of residence assume “yes” for the SA, the attacker will know that the victim has cancer. <u>Purpose</u>: It contrasts identity and attribute disclosure in background knowledge attacks. The metric is computed using *pyCANON* and *Anjana*.
- **δ-disclosure**: assuming our dataset has a single SA, let us define p(EC, s) as the fraction of patients in the EC with SA = s and p(DB, s) as the fraction of patients in the whole dataset (DB) with with SA = s. δ-disclosure is defined as^30^: 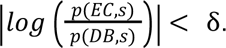 <u>Purpose</u>: it contrasts identity and attribute disclosure by ensuring SA prevalence in the dataset is below a threshold δ. The metric is computed using the package *pyCANON*.
- **basic β-likeness**: it restricts the relative maximal distance between the distribution of SAs in the EC and in the dataset. Let us define *P* as the distribution of SAs in the whole dataset DB and *Q* the distribution of SAs in a given EC, *basic β-likeness* is verified if 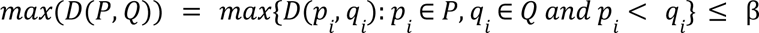 where 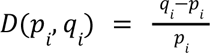 is the distance function between the two distributions^32^. <u>Purpose</u>: it contrasts attribute disclosure by limiting the concentration of SAs in ECs. The metric is computed using *pyCANON*.
- **t-closeness**: this metric requires that the distribution of a SA in any EC is close to the distribution of the attribute in the overall table, hence the distance between the two distributions should be no more than a threshold t^33^. A table is set to respect t-closeness if all EC in the table respect the threshold t. *t-closeness* is based on the Earth Mover Distance and it is bounded between 0 (no distance, hence high anonymity) and 1 (maximum distance, poor anonymity). <u>Purpose</u>: it contrasts attribute disclosure for skewness and similarity attacks. The metric is computed using the package *pyCANON* and *Anjana*.

### SDG algorithms

#### PrivBayes

PrivBayes is based on the construction of Bayesian networks, in which nodes are dataset features and link weights are their correlations^34^. The algorithm imputes a hierarchical relationship between nodes, establishing parent and children nodes, and estimates the conditional probability of observing a child feature from the parent feature in the original dataset. The user can set the maximum number of children nodes per parent. After this, the algorithm adds noise to the probability distributions to grant ε-differential privacy and generates the synthetic dataset from the hierarchic network.

#### PATEGAN

This algorithm is based on Generative Adversarial Networks (GAN), in which Private Aggregation of Teacher Ensembles (PATE) replaces the discriminator and ensures the differential privacy of the synthetic data^35^. A GAN consists of two neural networks, i.e. the generative and the discriminant network, playing a zero-sum game in which one’s gain corresponds to the other’s loss. The generative network is trained on the data to generate new data, possibly similar to the training set, hence minimizing the error between synthetic and original data thanks to a backpropagation process. The discriminant network is trained on the original and the synthetic data generated by the generative network. As more data is generated, the discriminant learns to discriminate better and better between synthetic and original data. GAN is a machine learning tool in which the generative network is trained to maximize the error of the discriminant network, i.e. to “trick” the discriminant network. Instead of using a discriminant network, the PATEGAN algorithm substitutes the discriminant process with a PATE process. The algorithm divides the dataset into *k* disjoint datasets and *k* classifiers (teachers) are trained on each of the subsets separately. For any given input vector, the algorithm classifies the vector through each of the *k* teachers, takes the most common output class among them, and adds Laplacian noise to the output to ensure ε-differential privacy.

#### TVAE

Similarly to GANs, variational auto-encoders (VAEs) are constituted by two neural networks, called the *encoder* and the *decoder*^36^. The encoder is trained on the original data to perform dimensionality reduction of the original features by projecting the data into a latent space of reduced features, while the decoder takes as input the data from the low dimensionality latent space and aims reconstructing the original data by minimizing the error. This version of VAEs overcomes the limitations of traditional VAEs that do not perform well with categorical variables. Tabular-VAE (TVAE) is generalized to manage both continuous and categorical variables, hence it applies to a vast range of data.

#### ADSGAN

ADSGAN (Anonymization through Data Synthesis using GANs)^37^ is a generative model designed to create synthetic datasets that closely approximate the statistical properties of original data while minimizing identifiability. The model builds upon conditional Generative Adversarial Networks and introduces a quantifiable definition of identifiability based on the probability of re-identification given all available patient data. By incorporating privacy-preserving mechanisms during training, ADSGAN aims to reduce the risk of data leakage while maintaining high fidelity.

#### Bayesian Network

The Bayesian Network approach for synthetic data generation is based on probabilistic graphical models, implemented within the pgmpy^38^ library used by synthcity. In this method, dataset features are represented as nodes in a Bayesian network, where directed edges indicate conditional dependencies between features. Conditional probability distributions are then estimated for each feature relationship. To enhance privacy, controlled noise is introduced into these distributions. Once the network is constructed, synthetic data is generated by sampling from the learned probability distributions.

### The Penta Platform

All our work was run on the Penta Platform, an AWS based cloud environment that allows users to connect their data with several computational workspaces such as RStudio, Jupyter Notebooks and more.

The Platform can be accessed by authorized users who can create their own studies. Studies are the users’ private directories of the platform, to which they can upload private data and decide whether to keep their data private or share them with a set of users. The Platform also provides a set of public data regarding several aspects of health.

Once the study has been defined, users are required to choose a workspace, which consists in defining the specs of the virtual machine in terms of power and storage. For our task we chose a virtual machine running with 64GB of RAM with GPU and 200GB of storage.

The virtual machine runs Anaconda, and is provided with multiple Jupyter environments, each of them associated with a Jupyter kernel. The set of pre-designed environments allow for the use of pytorch, Tensorflow and Spark, while a custom persistent environment allows the user to install any desired python packages, ensuring that the user will not have to reinstall the packages on the next login.

The user-friendly design of the machine ensures that the installation of packages on the custom environment and the workflow on the Jupyter notebook run as in a local terminal. For our work we used the custom persistent Kernel and we installed the packages synthcity^39^, synthgauge^40^, pyCANON^41^ and Anjana^42^.

### Synthetic Data Generation protocol

We tested state-of-the-art synthetic data generation (SDG) methods to generate new versions of available electronic health records (EHR) and test their capacity of reproducing previous results. In doing so, we also test the potential disclosure of sensitive private information present in the data, in order to grant the reproducibility of previous studies and foster further research use of the datasets in a privacy-compliant fashion.

We relied on the package *synthcity*^39^ to perform SDG and ran several tests to check statistical fidelity and the privacy level that the algorithms are able to preserve with respect to the original datasets. We employed five algorithms provided by the package *synthcity*, two that comply with differential privacy^28^, namely PrivBayes^34^, and PATEGAN^35^ and three, i.e. TVAE^43,44^, ADSGAN^37^ and Bayesian Networks^38^, that are not regulated by DP.

We integrated the employed algorithms, the chosen metrics and the performed tests in a protocol aimed at obtaining a synthetic dataset that preserves privacy requirements and ensures fidelity for the desired utility function. We outline the protocol in **Figure 1**.

**Figure 1:**
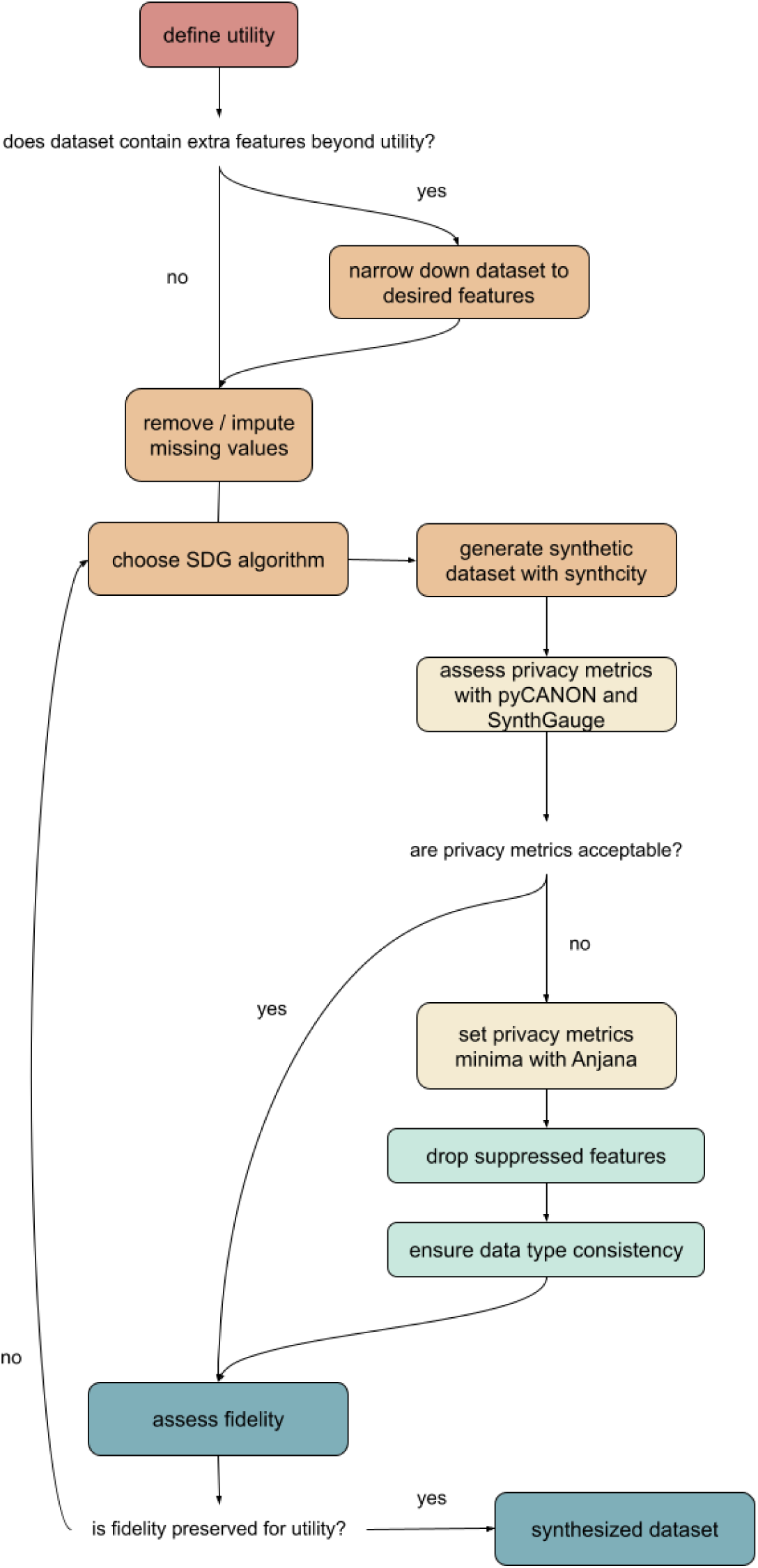
Flow diagram of the SDG protocol. We report the multiple steps necessary to complete the protocol of synthetic data generation for our cohort datasets.

As a first step, the data user must define the utility of the dataset, hence the scope of the data usage in the study. This may include estimating determinants, comparing time-series, realizing classification or other studies through statistical modeling or more sophisticated AI approaches. The user must discard the features that are not of interest, in order to maximise the potential of synthetic data generation, which is strongly hampered by high dimensionality. Data cleaning, as to remove null values from the tables or to perform missing values imputation, is critical to ensure seamless data generation. SDG algorithms can be chosen from the pool of available algorithms provided by *synthcity*. Here we focused on five algorithms, two of which ensure differential privacy, among others available. After realizing a batch of synthetic versions of the original dataset, the user must assess privacy metrics using *pyCANON* and *SynthGauge*. Easily, this first assessment will not satisfy all the privacy requirements. If this is the case, the user can rely on *Anjana*^42^ to correct the specific privacy metrics like k-anonymity, l-diversity and t-closeness by imposing minimum levels for these three metrics and a maximum suppression level, allowing for records suppression up to a maximum percentage. *Anjana* may suppress outliers and some features in order to reach the desired level of privacy. Removed or aggregated features may not allow for the full reproducibility of the original results, or may allow it in a different fashion: the user must check data type consistency. Finally, the user may assess the fidelity level of the final product, synthesized by *synthcity* and anonymized by *Anjana*, by reproducing the desider study and comparing it with the original study.

## Results

### Use case 1: Healthcare seeking behavior of influenza-like-illness digital cohorts

The first use case of SDG focuses on the determinants of healthcare seeking behavior for 6000 ILI episodes reported between 2011 and 2023 by 2184 individuals who voluntarily filled the InfluWeb^45^ online weekly survey with their information on symptoms severity, age, sex, educational level, working status, medical condition, duration of symptoms and belief on the cause of symptoms. Participants enrolled voluntarily to the study by registering on the Influweb website and receiving weekly reminders via email to fill in the survey.

We impose a *ε*-DP of 0.01. The *ε* parameter bounds the probability of finding the same output in two synthetic datasets generated from the input data, one with and one without a given user. This ensures that any user’s presence does not sensibly alter the output date, meaning that their presence in the original dataset cannot be easily inferred^28^.

In **Table 1** we observe the agreement between the odds-ratios measured on the original and a single version of the synthetic datasets for the two algorithms and their post-anonymization versions. Most significant (p-value < 0.05) predictors maintain significance both in the synthetic datasets and post-anonymized synthetic versions, except for a few variables highlighted in red. Infrequency of specific variables in the original dataset was preserved in the synthesized data. Missing odds-ratios correspond to infrequent variables in the synthetic datasets, corresponding to scarce variables in the original dataset, in most cases non-significant (p-value > 0.05). Notably, TVAE fails to reproduce small groups behavior, e.g. the “pregnancy” group, when their sample in the original data is small^46^ (30). Here we tested significance at 5%, inconsistencies are mostly found for predictors whose original significance was slightly stronger than 5%. Collecting wider samples, bootstrapping or testing for weaker significance may overcome these discrepancies. The opposite trend, i.e. post-synthesization acquired significance, is registered for pregnancy and higher education level. These appear to be significantly associated with higher healthcare seeking behavior, the former in the case of A-PrivBayes anonymized dataset, the latter in the TVAE synthetic dataset. These associations were not significant originally, however the significance of the original association was slightly weaker than 5% and generation noise may have altered it slightly. The association disappears in the anonymized A-TVAE version of the dataset, due to infrequency of the variable generated in the synthetic data.

**Table 1:**
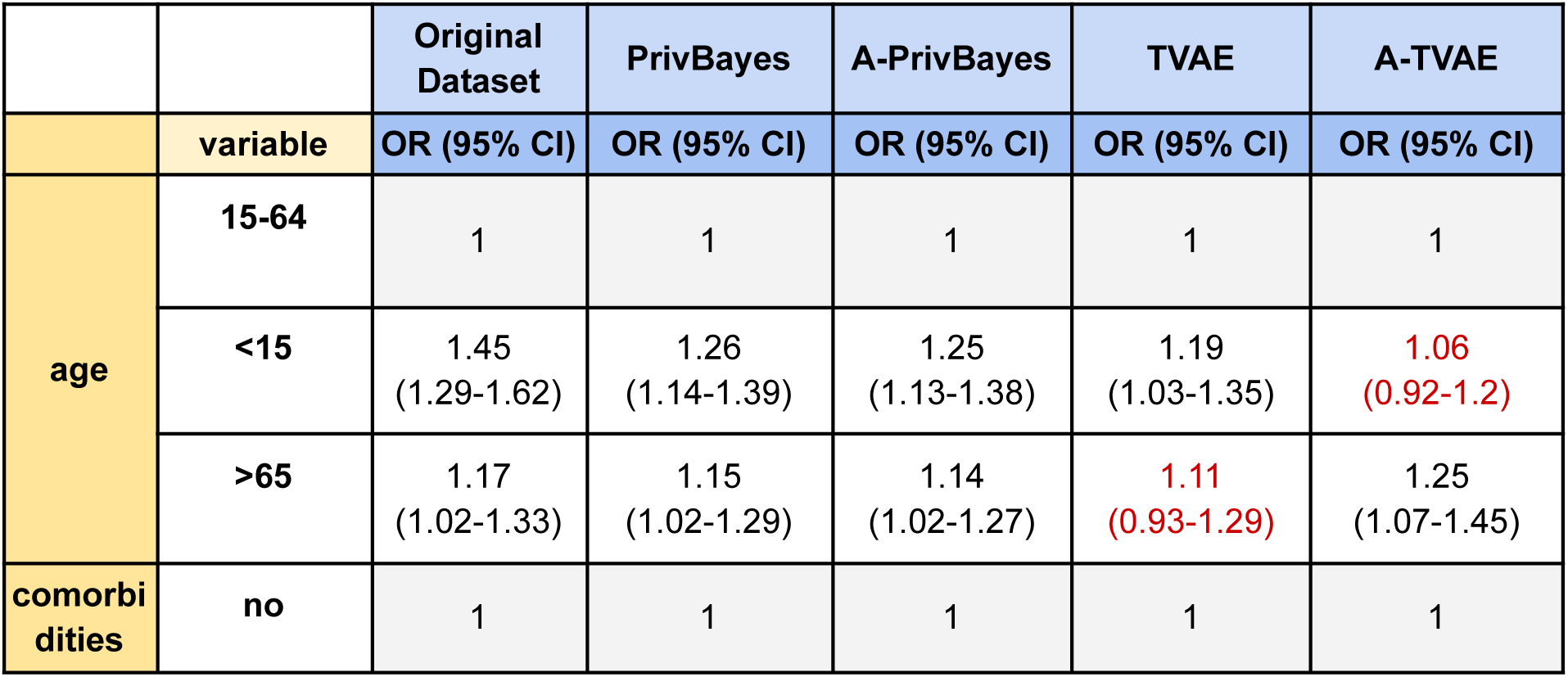
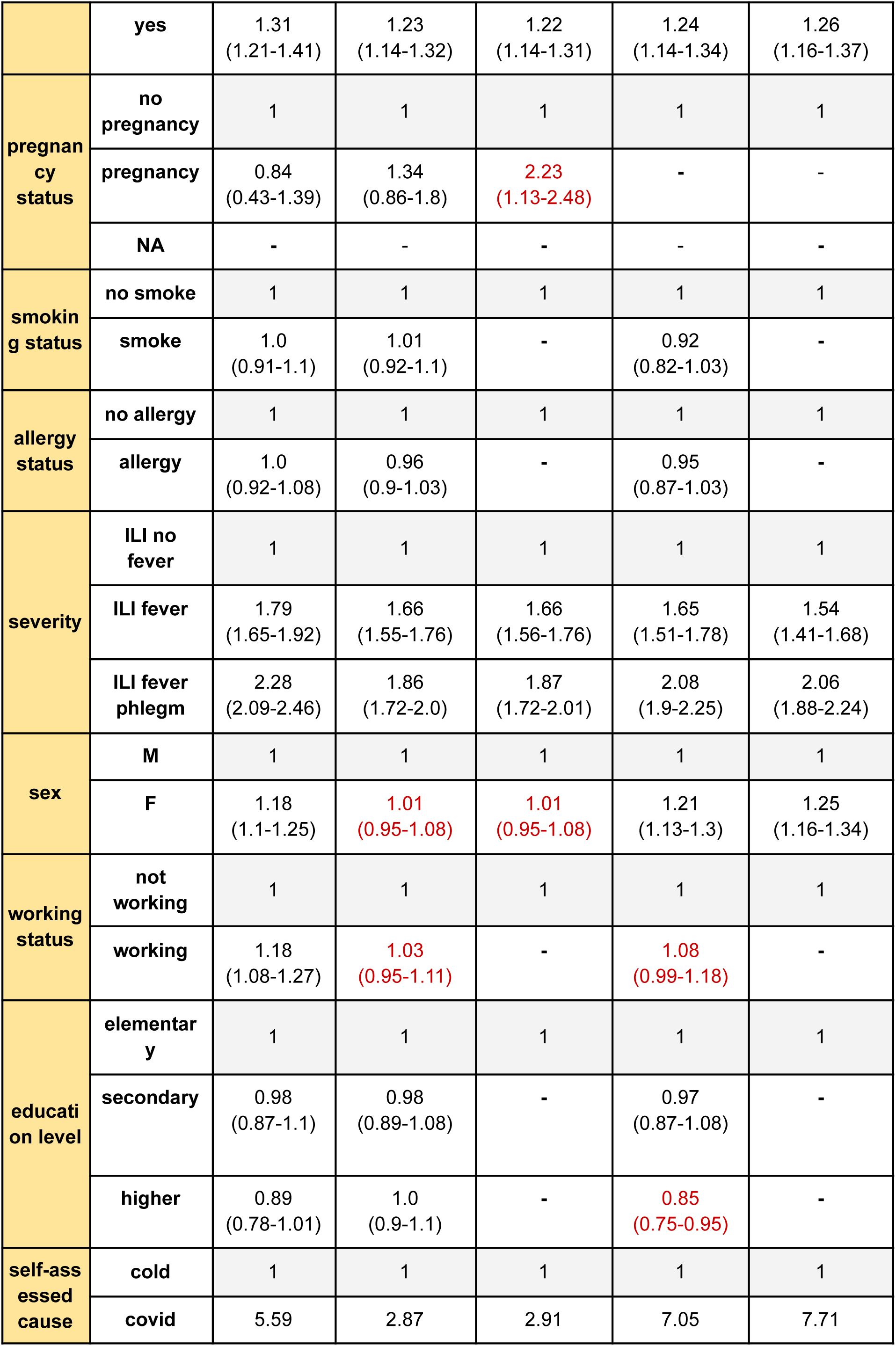
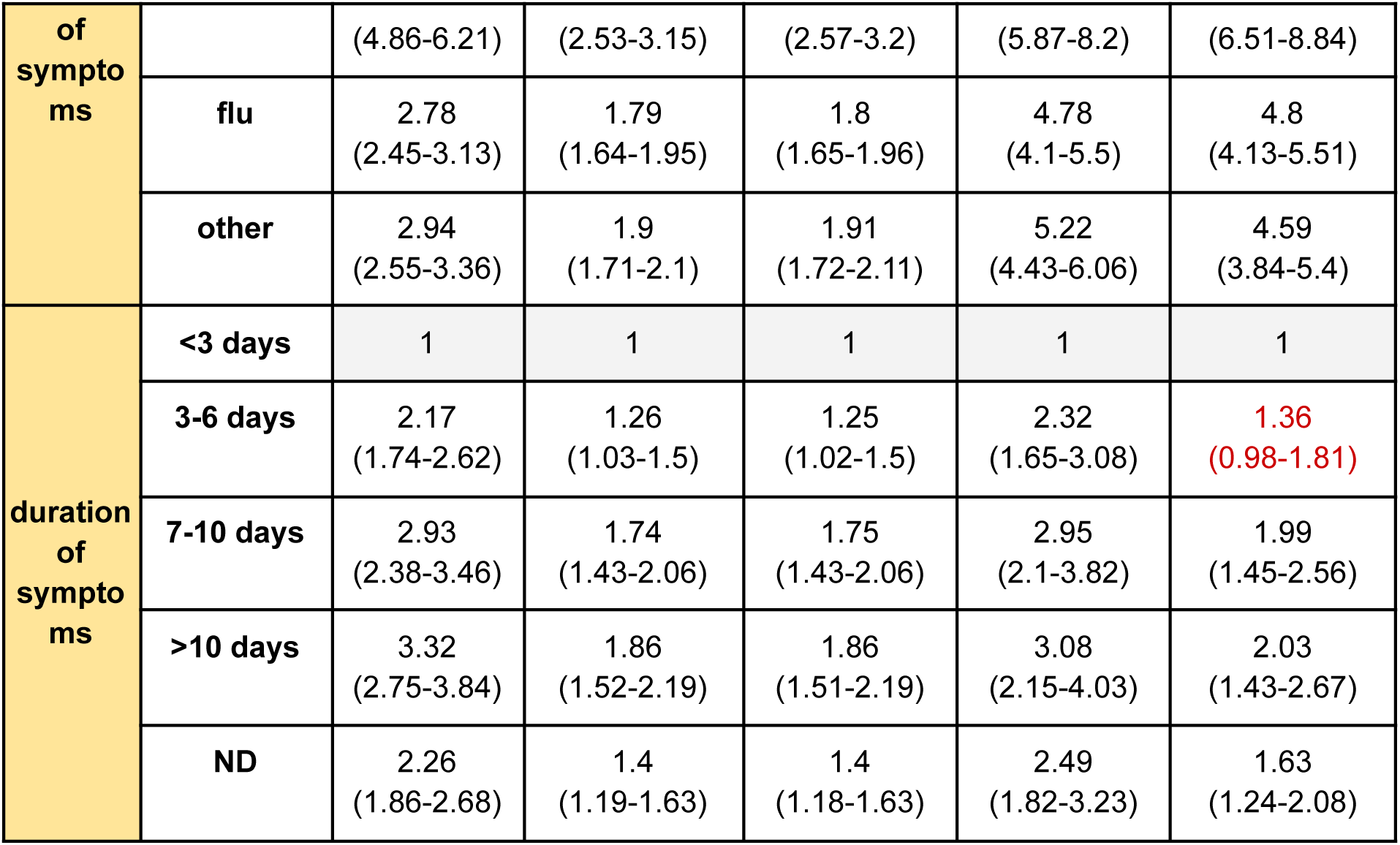
Logistic regression results comparison between the original and synthetic datasets generated by PrivBayes and TVAE. Estimates of healthcare seeking odds-ratios and 95% confidence intervals adjusted by the Zheng-Yu correction for outcomes that are more prevalent than 10%. Reference groups are color-coded in gray. Significance discrepancies are highlighted in red. Missing odds-ratios refer to infrequent variables.

We run some tests to compare the privacy preservation of the two algorithms relying on a set of benchmark privacy metrics from the anonymization literature.

For anonymized SDG algorithms like *A-PrivBayes* and *A-TVAE*, TCAP cannot be computed since *Anjana* removes identifiers. In **Table 2** we see how k-anonymity, l-diversity and t-closeness for *PrivBayes* and *TVAE* are not sufficiently high to avoid sensitive information disclosure. Anonymization through *Anjana* allows setting user-defined thresholds for these metrics, benefitting δ-disclosure and basic β-likeness as well, with less than 1% of the cohort data suppressed.

**Table 2:** Privacy metrics comparison. Metrics computed on 100 iterations of a synthesized dataset with 95% confidence interval. In yellow metrics computed with *SynthGauge*, in orange metrics with *Synthcity*, in red with *pyCANON*. We used “comorbidities” as SA, “age class”, “sex”, “working status”, “education level”, “pregnancy”, “smoking status” and “allergy status” as QIs and the users’ hashed-IDs as identifiers for TCAP. Values represent median and interquartile range of respective scores. Parameters used for Anjana, suppression level < 1%, *k-anonymity* > 5, *l-diversity* > 2, *t-closeness* < 0.5.

| Privacy metric | Reference | PrivBayes | A-PrivBayes | TVAE | A-TVAE |
| --- | --- | --- | --- | --- | --- |
| TCAP | - | 0.0001667<br>(0.0-0.0001667) | - | 0.0 (0.0-0.0002) | - |
| ε-DP | - | 0.01% | 0.01% | - | - |
| k-anonymity | 1 | 1.0 (1.0-1.0) | 9.5 (6.0-69.0) | 1.0 (1.0-1.0) | 21.0 (6.0-137.0) |
| l-diversity | 1 | 1.0 (1.0-1.0) | 2.0 (2.0-2.0) | 1.0 (1.0-1.0) | 2.0 (2.0-2.0) |
| δ-disclosure | 2.82 | 2.27 (2.05-2.61) | 0.67 (0.34-0.91) | 1.68 (1.62-1.86) | 1.22 (1.1-1.4) |
| basic β-likeness | 4.01 | 3.47 (3.41-3.56) | 0.48 (0.26-0.93) | 4.06 (3.95-4.14) | 1.9 (1.67-2.17) |
| t-closeness | 0.80 | 0.78 (0.77-0.78) | 0.13 (0.07-0.19) | 0.8 (0.8-0.81) | 0.37 (0.33-0.42) |

### Use case 2: Predicting 30 days post-admission mortality in hospital of patients diagnosed with sepsis

We leverage the MIMIC-III dataset^47,48^ comprising around forty thousands de-identified patients admitted to the Beth Israel Deaconess Medical Center in Boston, US, between 2001 and 2012, their time of admission and demission, diagnoses, demographics, vital signs, laboratory tests, health indicators such as blood pressure and heartbeat rate and mortality. Patients were deidentified, their sensible information removed and dates shifted to the future to preserve age at admission and demission, whereas patients over 80 appear as being 300 years old due to re-identification issues. Protected health information from text fields was removed. The project was approved by the Institutional Review Boards of Beth Israel Deaconess Medical Center (Boston, MA) and the Massachusetts Institute of Technology (Cambridge, MA). Requirement for individual patient consent was waived because the project did not impact clinical care and all protected health information was de-identified.

We replicate the analysis of a previous study predicting 30 days post-admission mortality of patients diagnosed with sepsis^49^. The study analyzed 4559 patients, separated in two groups based on patients’ survival after 30 days of hospital admission, resulting in 889 deaths and 3670 surviving patients. The work aimed at finding factors associated with mortality among their vital signs, trends, demographics and tests via a logistic regression. In **Table 3** we reproduce the results of the logistic regression for the original data and for two synthetic versions of the same dataset, generated with *PATEGAN* and *TVAE*.

**Table 3:** Logistic regression results comparison between the original and synthetic datasets generated by the PATEGAN and TVAE. In hospital 30 days post-admission mortality odds-ratios. Values represent the estimated median and 95% confidence interval of odds-ratios via logistic regression computed on the original datasets and their synthetic and post-anonymized synthetic versions. Discrepancies in significance are highlighted in red.

|  | Original Dataset | PATEGAN | A-PATEGAN | TVAE | A-TVAE |
| --- | --- | --- | --- | --- | --- |
| variable | OR (95%CI) | OR (95%CI) | OR (95%CI) | OR (95%CI) | OR (95%CI) |
| age | 1.03<br>(1.03-1.04) | 1.01<br>(1.0-1.01) | - | 1.01<br>(1.0-1.01) | - |
| aniongap_max | 1.03<br>(1.01-1.05) | 1.06<br>(1.05-1.08) | 1.03<br>(1.02-1.05) | 1.07<br>(1.05-1.08) | 1.06<br>(1.03-1.09) |
| bun_min | 1.02<br>(1.01-1.02) | 1.01<br>(1.0-1.01) | 1.01<br>(1.0-1.01) | 1.0<br>(1.0-1.01) | 1.01<br>(1.01-1.02) |
| creatinine_min | 0.73<br>(0.64-0.82) | 0.99<br>(0.95-1.03) | 1.02<br>(0.98-1.07) | 1.0<br>(0.96-1.04) | 0.94<br>(0.82-1.07) |
| heartrate_mean | 1.01<br>(1.01-1.02) | 1.01<br>(1.0-1.01) | 1.0<br>(1.0-1.01) | 1.01<br>(1.01-1.02) | 1.0<br>(1.0-1.01) |
| hematocrit_min | 1.0<br>(0.99-1.01) | 1.02<br>(1.0-1.03) | 1.02<br>(1.0-1.03) | 1.01<br>(1.0-1.02) | 1.02<br>(0.99-1.04) |
| lactate_min | 1.39<br>(1.28-1.52) | 1.1<br>(1.04-1.16) | 1.09<br>(1.04-1.15) | 1.08<br>(1.03-1.14) | 1.46<br>(1.31-1.62) |
| resprate_mean | 1.08<br>(1.06-1.1) | 1.1<br>(1.08-1.12) | 1.08<br>(1.06-1.1) | 1.08<br>(1.07-1.11) | 1.09<br>(1.05-1.12) |
| sodium_max | 1.0<br>(0.98-1.01) | 1.02<br>(1.01-1.04) | 1.02<br>(1.01-1.04) | 1.04<br>(1.02-1.06) | 1.06<br>(1.03-1.08) |
| sofa | 1.17<br>(1.14-1.21) | 1.08<br>(1.05-1.1) | 1.06<br>(1.04-1.09) | 1.08<br>(1.06-1.1) | 1.14<br>(1.1-1.19) |
| tempc_max | 0.73<br>(0.66-0.8) | 0.78<br>(0.72-0.86) | 0.77<br>(0.7-0.84) | 0.81<br>(0.74-0.89) | 0.57<br>(0.49-0.67) |
| vent | 1.79<br>(1.47-2.18) | 1.7<br>(1.45-2.0) | 1.46<br>(1.25-1.7) | 1.67<br>(1.43-1.96) | 1.28<br>(1.02-1.6) |
| wbc_max | 0.97<br>(0.95-0.98) | 1.0<br>(1.0-1.0) | 1.0<br>(1.0-1.0) | 1.0<br>(1.0-1.0) | 1.0<br>(0.98-1.02) |
| wbc_min | 1.07<br>(1.04-1.09) | 1.01<br>(1.0-1.01) | 1.01<br>(1.0-1.01) | 1.01<br>(1.0-1.01) | 1.02<br>(0.99-1.05) |

In **Table 3** we observe the agreement between odds-ratios computed on the original and synthetic datasets. Here again, most significant (p-value < 0.05) predictors maintain significance both in the synthetic datasets and in post-anonymized synthetic datasets, except for a few variables highlighted in red. Discrepancies in significance of predictors are mostly found for variables whose significance was slightly stronger than 5% threshold or those whose odds-ratios are strongly estimated as neutral (i.e. OR = 1), hence we do not interpret these as considerable differences. Age had a weak association with post-admission mortality. In the post-anonymized algorithms *A-PATEGAN* and *A-TVAE*, age is aggregated into age classes to meet privacy requirements. These age odds-ratios are not comparable with those computed on the original variable and we remove them for sake of comparison.

In **Table 4** we see how k-anonymity, l-diversity and t-closeness for *PATEGAN* and *TVAE* register too low scores to avoid sensitive information disclosure. Post-anonymization through *Anjana* allows for achieving user-defined values for these metrics, benefitting δ-disclosure and basic β-likeness as well, with less than 10% of the cohort data suppressed.

**Table 4:** Privacy metrics comparison. Metrics computed on 100 iterations of a synthesized dataset. In yellow metrics computed with SynthGauge, in orange metrics with *Synthcity*, in red with *pyCANON*. We used “mortality” as SA, “age” as QI and the users’ hashed-IDs as identifiers for *TCAP*. Values represent median and interquartile range of respective scores. Parameters used for Anjana, suppression level < 10%, *k-anonymity* > 10, *l-diversity* > 2, *t-closeness* < 0.5.

| Privacy metric | Reference | PATEGAN | A-PATEGAN | TVAE | A-TVAE |
| --- | --- | --- | --- | --- | --- |
| TCAP | - | 0.0 (0.0-0.0) | - | 0.0 (0.0-0.0002) | - |
| ε-DP | - | 5% | 5% | - | - |
| k-anonymity | 3 | 3.0 (2.0-4.0) | 11.0 (10.0-16.0) | 1.0 (1.0-1.0) | 46.5 (25.5-75.75) |
| l-diversity | 1 | 1.0 (1.0-1.0) | 2.0 (2.0-2.0) | 1.0 (1.0-1.0) | 2.0 (2.0-2.0) |
| δ-disclosure | 1.50 | 1.29 (1.08-1.46) | 1.07 (0.64-1.34) | 1.94 (1.67-2.18) | 0.81 (0.6-1.07) |
| basic β-likeness | 0.78 | 1.05 (0.76-1.26) | 0.64 (0.48-0.86) | 3.58 (2.94-7.49) | 0.45 (0.39-0.64) |
| t-closeness | 0.20 | 0.26 (0.23-0.29) | 0.18 (0.13-0.2) | 0.39 (0.32-0.88) | 0.07 (0.05-0.08) |

Similarly to the logistic regression case, in **Table 5** we see the statistical fidelity of the synthetic datasets with respect to the original data by comparing odds-ratios. Except for “creatinine min”, most odds-ratios are consistent across synthetic datasets. “creatinine min” appears positively associated with mortality in the *PATEGAN* synthetic dataset, while the association becomes non-significant (p-value > 0.05) in the *A-PATEGAN*, *TVAE* and *A-TVAE* datasets. Neutral odds-ratios in the original data may show fluctuations in the synthesized version, hence we do not interpret these as considerable discrepancies. Here as well, age had a weak association with post-admission mortality, which is captured by most of the synthetic datasets estimations. In the post-anonymized algorithm *A-TVAE,* age is anonymized into age classes, hence odds-ratios are not comparable with those computed on the original data.

**Table 5:** XGBoost regression results comparison between the original and synthetic datasets generated by the PATEGAN and TVAE. In hospital 30 days post-admission mortality odds-ratios. Values represent the estimated median and 95% confidence interval of XGBoost regression odds-ratios via bootstrap.

|  | Original Dataset | PATEGAN | A-PATEGAN | TVAE | A-TVAE |
| --- | --- | --- | --- | --- | --- |
| variable | OR (95%CI) | OR (95%CI) | OR (95%CI) | OR (95%CI) | OR (95%CI) |
| urineoutput | 1.0<br>(0.999-1.0) | 1.0<br>(1.0-1.0) | 1.0<br>(1.0-1.0) | 0.999<br>(0.999-0.999) | 0.999<br>(0.999-0.999) |
| lactate_min | 1.433<br>(1.347-1.543) | 1.163<br>(1.112-1.212) | 1.213<br>(1.163-1.279) | 1.324<br>(1.224-1.477) | 1.387<br>(1.292-1.503) |
| **bun_mean** | 1.016<br>(1.014-1.02) | 1.006<br>(1.003-1.008) | 1.008<br>(1.006-1.011) | 1.014<br>(1.01-1.018) | 1.014<br>(1.011-1.018) |
| **sysbp_min** | 0.989<br>(0.988-0.99) | 0.993<br>(0.992-0.994) | 0.992<br>(0.991-0.993) | 0.989<br>(0.988-0.99) | 0.99<br>(0.989-0.991) |
| **metastatic_cancer** | 2.761<br>(2.213-3.58) | 1.668<br>(1.35-2.232) | 2.523<br>(1.009-7.769) | 1.68<br>(1.067-2.379) | 4.609<br>(2.011-10.3) |
| **inr_max** | 1.067<br>(1.017-1.13) | 1.091<br>(1.059-1.13) | 1.12<br>(1.09-1.16) | 1.251<br>(1.15-1.376) | 1.254<br>(1.15-1.395) |
| **age** | 1.004<br>(1.002-1.006) | 1.004<br>(1.002-1.005) | 1.003<br>(1.001-1.005) | 1.006<br>(1.003-1.008) | - |
| **sodium_max** | 0.997<br>(0.997-0.998) | 0.997<br>(0.997-0.998) | 0.997<br>(0.997-0.997) | 0.996<br>(0.996-0.997) | 0.997<br>(0.996-0.997) |
| **aniongap_max** | 1.015<br>(1.008-1.022) | 1.007<br>(0.999-1.013) | 1.008<br>(1.001-1.015) | 1.021<br>(1.013-1.029) | 1.021<br>(1.012-1.028) |
| **creatinine_min** | 0.845<br>(0.792-0.896) | 1.107<br>(1.051-1.156) | 1.047<br>(0.991-1.092) | 0.967<br>(0.889-1.044) | 0.913<br>(0.86-1.009) |
| **spo2_mean** | 0.995<br>(0.994-0.995) | 0.995<br>(0.994-0.996) | 0.995<br>(0.994-0.996) | 0.994<br>(0.994-0.995) | 0.995<br>(0.994-0.995) |

Even in this use case, post-anonymization of synthesized datasets helps achieve higher privacy metrics by allowing suppression of less than 10% of the original data, as shown in **Table 6**. Suppressed data might contain outliers representing rare patients, which are a limitation for high privacy standards.

**Table 6:** Privacy metrics comparison. Metrics computed on 100 iterations of a synthesized dataset. In yellow metrics computed with *SynthGauge*, in orange metrics with *Synthcity*, in red with *pyCANON*. We used “mortality” as SA, “age” and “metastatic cancer” as QI and the users’ hashed-IDs as identifiers for *TCAP*. Values represent median and interquartile range of respective scores. Parameters used for Anjana, suppression level < 10%, *k-anonymity* > 10, *l-diversity* > 2, *t-closeness* < 0.5.

| Privacy metric | Reference | PATEGAN | A-PATEGAN | TVAE | A-TVAE |
| --- | --- | --- | --- | --- | --- |
| **TCAP** | - | 0.0 (0.0-0.0) | - | 0.0 (0.0-0.0002) | - |
| **ε-DP** | - | 5% | 5% | - | - |
| **k-anonymity** | 1 | 1.0 (1.0-1.0) | 10.0 (10.0-10.0) | 1.0 (1.0-1.0) | 14.5 (11.0-40.5) |
| **l-diversity** | 1 | 1.0 (1.0-1.0) | 2.0 (2.0-2.0) | 1.0 (1.0-1.0) | 2.0 (2.0-2.0) |
| **δ-disclosure** | 1.61 | 1.6 (1.51-1.77) | 1.58 (1.45-1.77) | 2.22 (2.12-2.31) | 1.21 (0.97-1.62) |
| **basic β-likeness** | 3.98 | 3.33 (3.24-3.42) | 1.43 (1.22-1.69) | 5.33 (5.21-5.51) | 0.8 (0.45-1.57) |
| **t-closeness** | 0.80 | 0.77 (0.76-0.77) | 0.32 (0.27-0.38) | 0.84 (0.84-0.85) | 0.14 (0.11-0.24) |

### Use case 3: Building a prediction model of in-hospital mortality in intensive care unit patients with heart failure

This exemplary use case is grounded on the results and methods of a reference research paper^50^, in which the authors built a machine learning model of in-hospital mortality of patients within the MIMIC-III dataset.

The reference dataset is of tabular format and comprises the data of 1177 heart failure patients with each row being dedicated to one patient. For each patient there are 49 features, as well as the final outcome value. The relevant outcome here is a true or false value, in-hospital mortality, which occurred for ∼13.52% of the patients. The features include basic values like age, biological sex or BMI, also existing preconditions like hypertension, atrial fibrillation or diabetes. Also included are bio measurements like heart rate, blood pressure and oxygen saturation, as well as laboratory values, like levels for lactic acid, anion gap or leucocytes.

First we cleaned and imputed missing values in the data, see Supplementary Material for the imputation methodology. We tested SDG performance with a range of built-in algorithms provided by *synthcity*, namely *Bayesian Network*, *ARF*, *ADSGAN*, *CTGAN*, *Normalizing Flows*, *PATEGAN*, *TVAE*, *RTVAE*, *PrivBayes*, *GOGGLE*, *GreaT*, *DPGAN*, and *DECAF*. Data generation was conducted using the recommended default parameters with encoder noise scale set to 0.1 for *Bayesian Network*. We restricted our focus on the twenty features studied in the reference paper, which represent the utility of our synthetic datasets. Through a multi-step process (see **Supplementary Figure SI-1**), we determined the two best performing algorithms that applied for the given use case. These are *Bayesian Network* and *ADSGAN*, as they performed best at reproducing single-variable marginals and statistical relationships among variables.

We assessed privacy metrics after the first synthesization and, in order to match standard privacy levels, the datasets were further anonymized using *Anjana*. Finally, we assessed the statistical similarity of the final datasets by reproducing the original analysis and comparing outcomes produced on the resulting synthetic and original datasets.

Figure 2 illustrates the agreement on the relationship between the risk of in-hospital mortality and four predictors of mortality highlighted in the original study with those reproduced from our synthetic datasets. The four features exhibit strong correlations in both the reference dataset and the synthetically generated datasets. This high statistical resemblance suggests that the synthetic data maintains sufficient quality for replicating critical findings and preserves utility.

**Figure 2:**
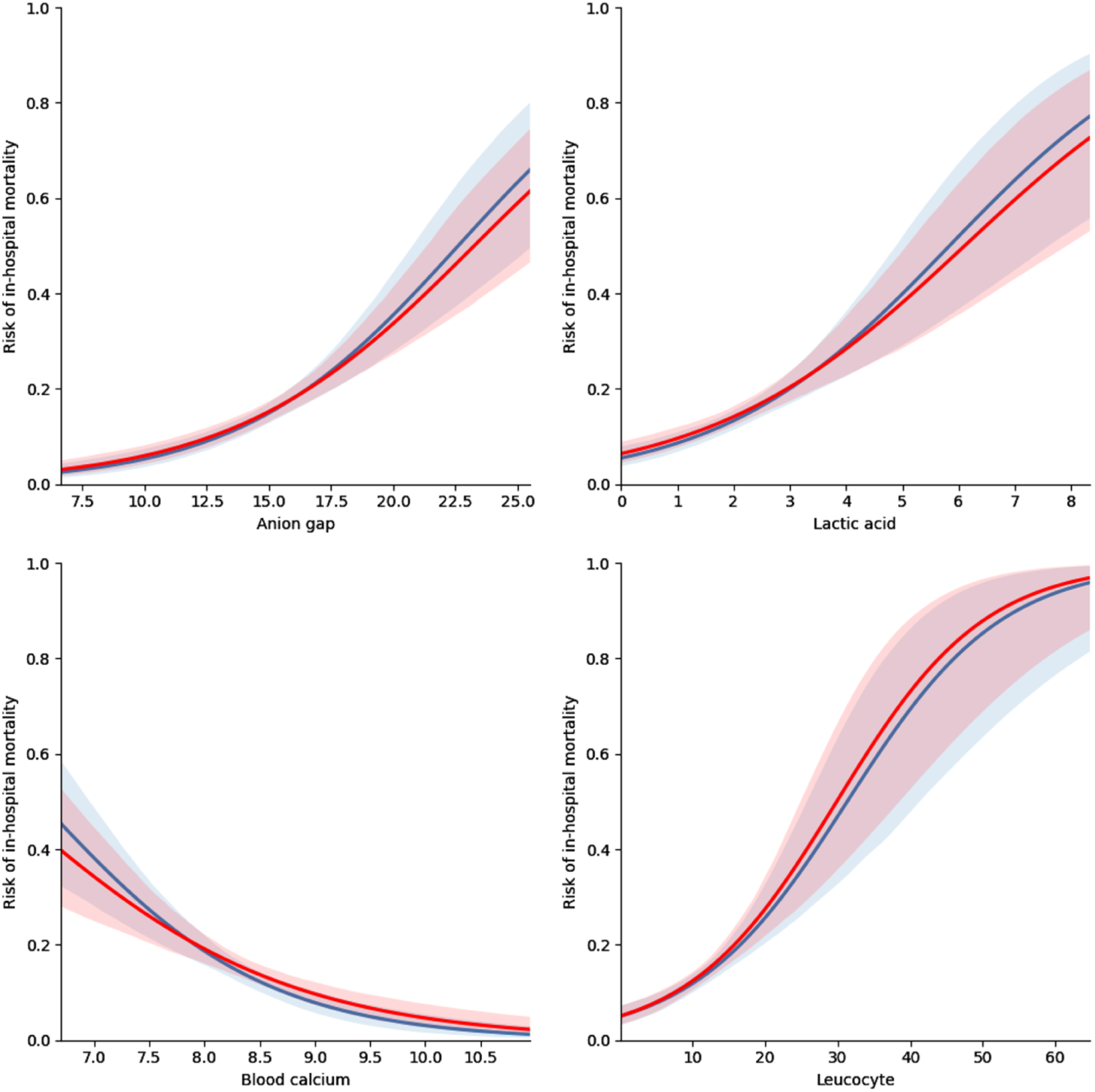
Comparison of associations between in-hospital mortality with four predictors estimated from the reference dataset and synthetic dataset. The logistic regression plots and 95% CI in blue stem from the reference dataset, while the red plots were derived from the dataset created with the Bayesian Network method after post-anonymization with *Anjana*.

Assessing privacy in synthetic datasets is inherently challenging. A direct inspection for “leaked rows”—exact copies of real data—revealed none. To further evaluate privacy risks, we computed several privacy metrics, comparing the synthetic dataset with the original using *pyCANON*. These included: k-anonymity, l-diversity, δ-disclosure, basic β-likeness and t-closeness. For all metrics, we used the outcome as the SA (mortality), the patient age and the categorical feature “Renal failure” were used as quasi-identifiers. For reidentification measures, the unique ID was the identification target.

The resulting privacy measurements are comparable to those of the reference dataset, suggesting that privacy characteristics were largely similar (see **Table 7**). Additionally, using *SynthGauge*, the T-CAP score was calculated to be 0, which suggests a low risk of individual traceability.

**Table 7:** Privacy metrics compared between reference dataset and synthetic versions. Metrics computed on 100 synthesized and Anjana-anonymized datasets, as well as the reference dataset. In yellow are metrics computed with *SynthGauge*, in red with *pyCANON*. We used “mortality” as SA, “age” and “renal failure” as QI and the users’ hashed-IDs as identifiers for *TCAP*. Values represent median and interquartile range of respective scores. Parameters used for *Anjana*, suppression level < 10%, *k-anonymity* > 10, *l-diversity* > 2, *t-closeness* < 0.5.

| Privacy metric | Reference | Bayesian Network | A-Bayesian Network | ADSGAN | A-ADSGAN |
| --- | --- | --- | --- | --- | --- |
| **TCAP** | - | 0.0 (0.0 - 0.0) | - | 0.0 (0.0 - 0.0) | - |
| **k-anonymity** | 1.0 | 1 (1.0 - 1.0) | 13 (11.0-19.0) | 1 (1.0 - 1.0) | 13 (11.0 - 43.25) |
| **l-diversity** | 1.0 | 1 (1.0 - 1.0) | 2 (2.0-2.0) | 1 (1.0 - 1.0) | 2 (2.0 - 2.0) |
| **δ-disclosure** | 2.0 | 2.0 (1.96 - 2.04) | 1.14 (0.97 - 1.61) | 2.01 (1.93 - 2.07) | 0.75 (0.49 - 1.08) |
| **basic β-likeness** | 6,4 | 6.4 (6.08 - 6.69) | 0.98 (0.48 - 1.07) | 6.44 (5.91 - 6.95) | 0.46 (0.3 - 0.72) |
| **t-closeness** | 0.86 | 0.86 (0.86 - 0.87) | 0.13 (0.08 - 0.16) | 0.87 (0.86 - 0.87) | 0.08 (0.06 - 0.10) |

Next, the privacy levels were heightened further through post-anonymization. We configured *Anjana* to ensure a k-anonymity of at least 10, l-diversity of at least 2 and t-closeness below 0.5. We set the suppression limit to a maximum of 10% rows. It should be mentioned at this point that both *Bayesian Network* and *ADSGAN* provide configurable privacy settings during the generation process, allowing for further control over privacy levels. So if privacy is still deemed too low, the generation process can also be tuned to improve privacy levels this way, but this is also likely to impact data resemblance, again highlighting the aforementioned trade-off.

In the post-anonymization step the “age” QI aggregation was modified to enhance privacy preservation and meet the user-defined requirements. For the dataset generated with the *Bayesian Network* algorithm, age values were grouped into 5-year intervals, whereas in the *ADSGAN*-generated dataset, they were aggregated into 10-year spans. Notably, *Anjana* did not require censoring of the “Renal failure” feature in either case.

The privacy metrics calculated on the post-anonymized datasets indicate a substantial improvement, as all evaluated metrics showed considerable enhancement. Despite these modifications, data resemblance remained largely unaffected. The only directly transformed feature, “age,” retained sufficient granularity to preserve the key relationships observed in the cleaned reference dataset (see **Figure SI-2**). Additionally, only ∼2.6% of the rows had to be removed in the *Bayesian Network* dataset, minimizing data loss while strengthening privacy.

## Discussion

In this work we designed a two-step protocol applicable to both continuous and categorical panel data based on four public and off-the-shelf packages of SDG (*synthcity*), privacy assessment (*pyCANON* and *SynthGauge*) and anonymization (*Anjana*). By using this protocol, the data owner is able to generate, assess and correct the privacy levels of their synthetic datasets based on specific metrics like k-anonymity, l-diversity and t-closeness before releasing a version for public use. This potentially opens the doors to renewed praxis in data governance^7^ allowing reutilization of past collected electronic health records like hospital admissions and cohort studies. Such policies could boost data sharing into open and secure frameworks like the European Health Data Space for data collections financed by public funding. This would strengthen European data readiness, helping the community access a common data space of shared records collected by concurrent research projects.

We employed several of the most common privacy metrics in our study, however no unique solution provides full protection from all attacks and a combination of measures is needed to minimise vulnerabilities. DP is not a general solution for maintaining individual-level records privacy. DP is a composable metric, meaning that accessing multiple releases of a synthetic dataset does not help the attacker gain more information on the original identities. However, the presence of outliers or leaked records from the original data exposes DP processes to linkage attacks, like membership inference and attribute inference^5^. For this reason we employed common anonymization metrics like k-anonymity and l-diversity to monitor outliers and minimize re-identification and attribute disclosure risks. Post-anonymization with *Anjana* ensures that outliers are suppressed in order to comply with privacy requirements.

A further weakness of DP is that DP-processes do not perform well on small datasets, which are common for cohort studies on rare diseases. Small datasets provide low utility for DP processes, since having smaller populations translates into “harder to hide” contributions of each individual in the dataset^4^, hence lowering utility. In small datasets, fidelity is affected by randomization applied to the data by the DP-process to match the maximum privacy loss.

Studies have shown that increasing the scale of data improves the signal-noise ratio^4,51^, formally referred to the scale-epsilon exchangeability. Hence, collecting more data allows to achieve the same utility preserved by DP with a smaller dataset at the cost of a lower privacy loss^4,28,52^. However, this might not always be possible and it does not come without downsides. Collecting more data has a disparate effect on the fidelity retained for underrepresented populations with respect to more populated groups^53^. This makes DP less supportive of studies on rare diseases or minorities when part of a larger and more general cohort.

k-anonymity is not composable and it is vulnerable to linkage attacks if not combined with l-diversity^31^ and t-closeness^33^. This implies that SDG should allow limited releases of synthetic datasets to avoid that privacy levels matched by the protocol are actually releasing more information that desired, i.e. two k-anonymous datasets synthesized from the same original dataset might be not k-anonymous^27^.

Outliers not only expose DP SDG algorithms to linkage attacks, they also affect k-anonymity. Post-anonymization with *Anjana* ensures that outliers are removed to avoid this vulnerability.

However, this implies that SDG is not a viable solution to support analyses on rare events and underrepresented populations.

We proved that synthetic data generation is a sound and viable solution to grant reproducibility of previous studies with high statistical fidelity, even with datasets of moderate data dimensionality, i.e. ten to twelve dimensions, both categorical or numerical, from cross-sectional or longitudinal studies. EHR are typically high dimensional by design of the data collection study and this strongly limits anonymization metrics^54^. In some cases high dimensionality of the collected dataset is due to the extensive number of known determinants of the health outcome under study, in other cases this is due to collecting as much data as possible as to widen the window of research for multiple hypotheses testing. Due to this, achieving high levels of privacy for synthetic data generation is not trivial due to the high dimensionality of most of the EHR collected. In synthetic data generation, high dimensionality imposes a narrow trade-off between high statistical fidelity of the generated dataset with respect to the original data and privacy preservation of sensible features^5^. In our protocol we imposed a process of dimensionality reduction that is context dependent, in which the user must narrow down the dataset features to the essential set to maintain utility. In our case studies, we also tested the processing of a wider number of features from the original datasets. In all cases, doing so led to increased risk of sensible information disclosure or loss of statistical fidelity. This tradeoff has been already observed by previous studies and comes from mathematical limitations imposed by the privacy metrics definition. Intuitively, datasets with a high number of sensitive variables are more exposed to re-identification. In this scenario, imposing high statistical fidelity at the cost of privacy leads to a higher risk of rare individuals (outliers) leaking to the synthetic dataset, hence recovering the vulnerabilities explained above. Similarly, the high number of sensitive features imposes low k-anonymity and l-diversity, by definition of EC, hence exposing the data to information disclosure. On the other hand, imposing high privacy preservation at the cost of statistical fidelity is useless from the point of view of utility for the research itself.

Algorithms ensuring differential privacy generally perform better at maintaining high levels of privacy, our two-step protocol of synthetization and post-anonymization allows for the use of a wider pletore of algorithms, ensuring outliers’ suppression and performing better on statistical fidelity, without concern for information disclosure.

In all cases, synthetic datasets must be handled with care. Here we provided a systematic analysis of important privacy metrics, assessed the reproducibility of past studies and showed how fidelity is preserved by a single realization of each SDG algorithm employed. However, results may vary across multiple realizations and safety checks on statistical fidelity are always needed before any further analysis or data release. Most algorithms are based on stochastic processes and sampling, hence safety checks ensuring high standards of fidelity in all realizations are needed, as highlighted at the end of the protocol. All in all, the recipe for a good synthetic data generation cannot prescind from a parsimonious features selection that allow our protocol to perform best on both fidelity and privacy outcomes.

## Conclusion

Synthetic data generation is an emerging technique that proved useful to ensure reproducibility of past studies and should be encouraged by public institutions to broaden the spectrum of studies for the health research community.

Our two-step protocol applies to a wide range of panel data, allowing data owners to generate and share high privacy - high fidelity synthetic datasets for public use, which could not be shared otherwise due to privacy restrictions.

Good SDG protocols cannot prescind from parsimonious features selection, minimising at all costs high data dimensionality, which poses a limitation to the use of SDG techniques for exploratory studies.

Data governance policies adopting SDG in publicly funded research may strongly enhance reutilization of data from historic cohort studies, enriching the European Health Data Space and strengthening data readiness for future health threats.

## Limitations

SDG proved useful to preserve high levels of privacy in simulated data with a good statistical fidelity of observed relations between features. However, we could not maintain both fidelity and privacy levels to the maximum for high dimensionality datasets. Reproducibility of previous results was limited to the last mile of analysis, hence not covering the entire process of data cleaning involving high dimensional datasets.

Data suppression in the final anonymization steps ensures that user-defined requirements for privacy metrics are met, at the cost of outlier suppression. Outliers could represent important data points that in new explorative research may drive to new results and new research lines. However, these data points are incompatible with privacy preservation standards and cannot be saved by SDG.

## Data availability

The Influweb dataset analysed during the current study is not publicly available due to privacy and confidentiality standards.

The MIMIC-III dataset analysed during the current study is publicly available at https://physionet.org/content/mimiciii/1.4/ upon free registration and course completion. The use of the database for research purposes is granted to all researchers provided that they take an online course and pass a test on ethics of human subjects research and experiments, achieve certification from the MIT and sign a Data Usage Agreement. The authors M.M. and J.E. passed the test, signed the DUA and were granted access to extract data from this database for research purposes. Extracted datasets for use cases 2 and 3 are also publicly available at the URLs of the respective studies.

The synthetic Influweb dataset generated during the current study will be available upon acceptance at the Zenodo repository, [persistent URL to appear here].

## Code availability

The underlying code used in this study for generating the synthetic data in three use cases will be made public upon acceptance in a GitHub repository and will be accessible via this link [URL to appear here].

## List of legends

ADSGAN: Anonymization through Data Synthesis using Generative Adversarial Networks
BMI: Body Mass Index
DP: Differential Privacy
EC: Equivalence Class
EHR: Electronic Health Records
GDPR: General Data Protection Regulation
HIPAA: Health Insurance Portability and Accountability Act
ILI: Influenza-Like-Illness
IRB: Institutional Review Board
PATEGAN: Private Aggregation of Teacher Ensembles Generative Adversarial Networks
QI: Quasi-Identifier
SA: Sensitive Attribute
SDG: Synthetic Data Generation
TCAP: Targeted Correct Attribute Probability
TVAE: Tabular Variational Auto-Encoders

## Conflict of interest

All authors declare no financial or non-financial competing interests.

## Author contributions

M.M.: Conceptualization, methodology, data curation, analysis, writing original draft and editing.

J.E.: Methodology, data curation, analysis, writing original draft and editing.

P.S.: Software curation and editing.

M.H.: Supervision, funding acquisition and editing.

M.D.T.: Project administration, conceptualization, supervision, funding acquisition, writing and editing.

D.P.: Project administration, conceptualization, supervision, funding acquisition, writing and editing.

## Acknowledgements

M.M. and D.P. acknowledges support from the Lagrange Project of the ISI Foundation, funded by Fondazione CRT. M.M. acknowledges support from the EOSC project SIESTA (101131957) funded by the European Union’s Horizon Europe research and innovation programme. All the authors acknowledge support from the Horizon Europe project VERDI (101045989), funded by the European Union. The funders played no role in study design, data collection, analysis and interpretation of data, or the writing of this manuscript. Views and opinions expressed are those of the author(s) only and do not necessarily reflect those of the European Union or the European Health and Digital Executive Agency. Neither the European Union nor the granting authority can be held responsible for them.

## Ethical statement

For Influweb the research was conducted in agreement with Italian regulations on privacy and data collection and treatment. The institutional review board of ISI Foundation, upon consultation with the Italian Data Protection authority, waived the ethical approval for the study.

## Notes

### Competing Interest Statement

The authors have declared no competing interest.

