## Supplementary Information for "Assessing synthetic data generation utility for cohort data secondary use"

### Descriptive statistics for use case 1

| | variable | fraction % (occurrence) |
|---|---|---|
| healthcare seeking | no | 61.05 (3663) |
| | yes | 38.95 (2337) |
| age | 15-64 | 82.55 (4953) |
| | <15 | 9.4 (564) |
| | >65 | 8.05 (483) |
| sex | F | 48.88 (2933) |
| | M | 51.12 (3067) |
| working status | not working | 42.85 (2571) |
| | working | 57.15 (3429) |
| education level | elementary | 21.83 (1310) |
| | higher | 42.4 (2544) |
| | secondary | 35.77 (2146) |
| duration of symptoms | <3 days | 4.78 (287) |
| | 3-6 days | 13.77 (826) |
| | 7-10 days | 4.02 (241) |
| | >10 days | 3.33 (200) |
| | ND | 74.1 (4446) |
| comorbidities | yes | 19.98 (1199) |
| | no | 80.02 (4801) |
| pregnancy status | no pregnancy | 99.48 (5969) |
| | pregnancy | 0.52 (31) |
| smoking status | not smoking | 84.33 (5060) |
| | smoking | 15.67 (940) |
| allergy status | allergy | 32.53 (1952) |
| | no allergy | 67.47 (4048) |

| | | |
|---|---|---|
| **self-assessed cause of symptoms** | **cold** | 19.8 (1188) |
| | **covid** | 2.68 (161) |
| | **flu** | 61.32 (3679) |
| | **other** | 16.2 (972) |
| **severity** | **ILI no fever** | 48.33 (2900) |
| | **ILI fever** | 39.28 (2357) |
| | **ILI fever phlegm** | 12.38 (743) |

**Table SI-1: Descriptive statistics of the dataset for use case 1.** Fraction and occurrence of ILI episodes for all features present in the dataset.

#### Imputation procedure for use cases 3

Initially, missing values were identified, and an appropriate representation of the existing data distribution was determined. The distribution was then estimated and used to generate plausible replacements for the missing entries. Imputation was performed using one of three approaches: normal distribution, lognormal distribution, or median substitution, depending on the characteristics of the observed data. To ensure the validity of the imputation process, histograms of the dataset before and after imputation were visually compared, confirming that no obvious bias was introduced.

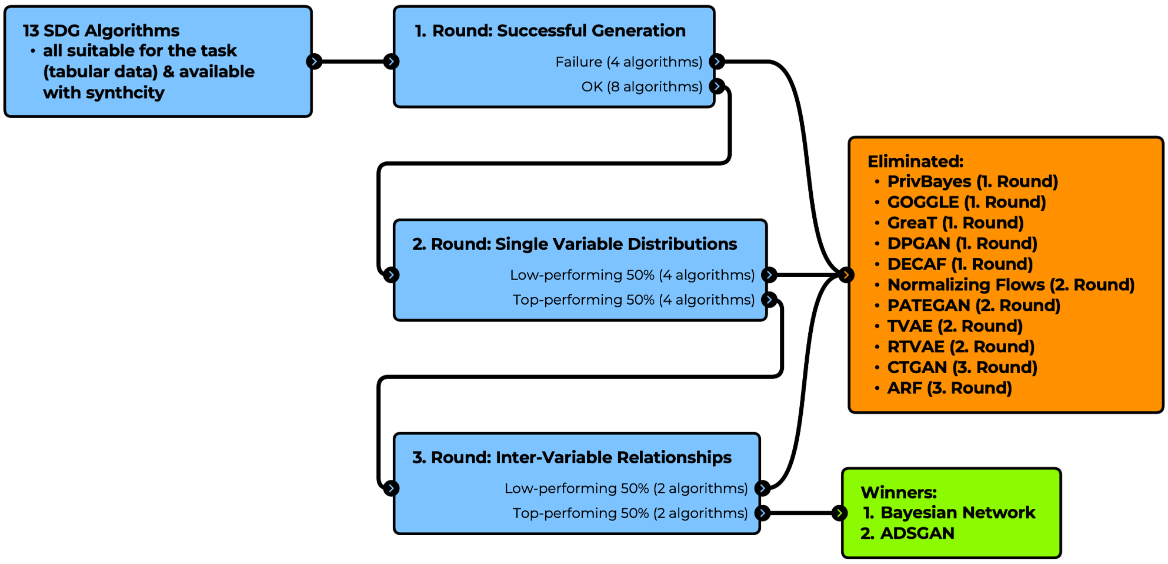


**Figure SI-1: Overview Benchmark Process.** A multi-step process used to select the best performing SDG algorithm for the reference dataset used in use case 3.

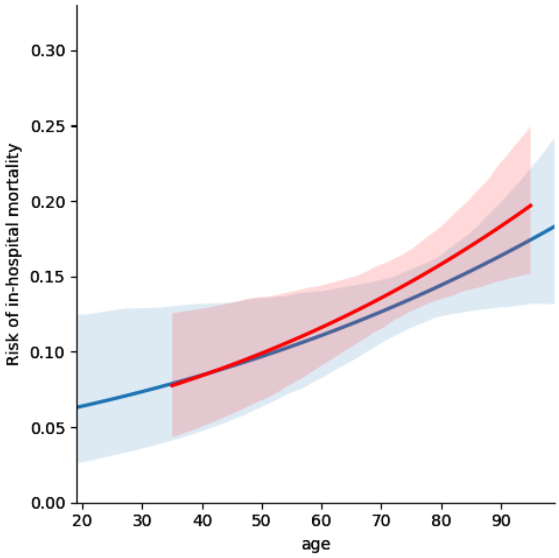


**Figure SI-2: Comparison of inter-variable relationship of patient age modeled within reference dataset and synthetic dataset after Anjana anonymization.** The logistic regression plots and 95% CI in blue stem from the cleaned reference dataset, while the red plots were derived from the dataset created with the Bayesian Network method after Anjana anonymization. The relationships appear to have been captured correctly and were reproduced well within the synthetic dataset, even after categorization of the age values.
